# Evidence that intergenerational income mobility is the strongest predictor of drug overdose deaths in U. S. Heartland counties

**DOI:** 10.1101/2023.07.18.23292832

**Authors:** Gene M. Heyman, Ehri Ryu, Hiram Brownell

**Author notes:** Corresponding author. (GMH).

## Abstract

In 2017, the Acting U. S. Secretary of Health and Human Services declared the “opioid crisis” a nation-wide health emergency. However, the crisis’s geography was not nation-wide. Many counties and towns had no overdose deaths, whereas others were home to hundreds. According to many influential research reports and news stories, geographic variation in overdose deaths was due to geographic variation in opioid prescription rates and/or geographic variation in socioeconomic factors, such as unemployment. Our goal was to test the degree to which prescription rates and socioeconomic correlates of income inequality predicted overdose deaths in the 1055 U.S. Midwest (“Heartland”) counties over the years 2006 to 2020. We used multilevel regression models to gauge the predictive strength of overdose rates and six socioeconomic measures that are correlated with income inequality. There were significant state-level and county-level differences. Intergenerational income mobility was the strongest predictor of overdose deaths, with regression coefficients that averaged about twice as large as the coefficients for opioid prescription rates. Every year, counties with greater upward intergenerational income mobility had lower overdose death rates. Social capital had the second largest regression coefficients, albeit by a small margin. Counties are the smallest demographic unit for which drug overdose rates are available; the results of this study link growing income inequality and drug overdose deaths at the county level.

## Introduction

In 2013 the U.S. Department of Health and Human Services warned that prescription opioid misuse had reached epidemic levels; in October of 2017, President Trump declared drug use, addiction, and opioid overdoses a national health emergency. The declarations were in response to dramatic increases in the availability of prescription opioids and the concomitant rise in drug overdose deaths. The amount of legally prescribed opioids, as measured in morphine equivalent grams, was three times greater in 2013 than in 2001; similarly, the number of drug related overdose deaths was about three times greater in 2013 than in 2001 [1, 2]. These changes are at the national level; at a more granular level, higher prescription rates were correlated with higher overdose rates in states, counties, and individuals [3-5]. Accordingly, many researchers and journalists explained the increases in overdose deaths in terms of the increases in legal opioid prescriptions [6-10]. State legislators translated these observations into prescription monitoring programs that had the goal of reducing the number of prescriptions that physicians wrote and, thereby, reducing overdose deaths By 2019, every state in the country but Missouri had an opioid prescription monitoring program in place [11]. Nevertheless, as some researchers and journalists pointed out, there were good reasons to believe that socioeconomic factors also played an important role in the opioid crisis [12-16]. However, the socioeconomic factors that are most critical have not been established, and, accordingly, their importance relative to prescription rates remains unknown. Our goal was to address these issues at the county level. Counties are the smallest geographic unit for which overdose data are available, and regression coefficients were our measure of importance.

### The opioid crisis is one of a series of linked drug epidemics

Two, well-established observations support the hypothesis that socioeconomic factors are critical to understanding the opioid crisis. First, from 2013 to the present, opioid prescription rates have steadily declined, yet drug overdose rates have continued to increase and at slightly higher rates than prior to 2013. That is, over the past ten years or so, opioid prescription and drug overdose rates have moved in opposite directions. The rising death toll is due to the in increased use of two illegal drugs: heroin and fentanyl [17, 18]. This transition is not unique, but one of several that have taken place since at least 1978, according to an important analysis by Jalal and colleagues [19]. Their graphs reveal that overdose death rates have been increasing at a steady rate for at least the past 45 years or so, counting all drugs, not just opioids. That is, the “opioid epidemic” is one of a series of “subepidemics” [19]

Second, history shows that large scale changes in the use and misuse of drugs, such as the opioid crisis, are often associated with large scale demographic factors. For example, when heroin was legal in the U. S., its use was not widespread, but limited to large East Coast cities [20]. Then, when legal access to opiates was restricted (the 1914 Harrison Narcotics Tax Act), decreases in their use varied according to social class, occupation, and related characteristics [21]. This is a familiar story: everyday observation reveals demographic correlates of alcohol, cigarette, and marijuana use. Taken together, Jalal et al.’s analysis and the history of American drug use, suggest that the “opioid epidemic” is one phase of an on-going and long-standing overdose trend that has strong ties with socioeconomic factors as well as with increased access to legal opioids by way of prescriptions.

### The relative importance of prescriptions and socioeconomic factors

Researchers have explored the role that socioeconomic factors might play in opioid overdose deaths, but no clear pattern has emerged. Ruhm [22] found that counties that had experienced economic downturns had higher overdose rates, but he concluded that county differences were explained better by the price and availability of drugs. In contrast, Zoorob and Salemi [23] tracked Medicare opioid prescription rates and concluded that social capital [24] provided the stronger account, titling their report: “Bowling alone, dying together.” However, in these studies, the research approach may have limited the findings. Zoorob and Salemi used Medicare prescriptions to measure opioid availability, but most Medicare patients are 65 and older. Ruhm used familiar macroeconomic variables such as poverty rate, income, home price and unemployment, but these may not adequately capture the conditions that motivate drug use.

### Our approach

We used multilevel multiple regression to quantify the degree to which drug availability and intergenerational income mobility explain county-level variation in drug overdose deaths from 2006 to 2020 in the 1055 counties of the twelve Midwest states: Illinois, Indiana, Iowa, Kansas, Michigan, Minnesota, Missouri, Nebraska, North Dakota, Ohio, South Dakota, and Wisconsin. Midwest overdose trends are representative of U.S. national trends, and as described in the *Discussion* section of this paper, previous research indicates that Midwest counties will provide a clearer account of the relations between prescriptions, intergenerational income mobility and overdoses than will other regions of the country. Our approach was based on our previous state-level study [5], which, in turn, was based on the parallels between geographic variation in overdose deaths and Chetty et al.’s [25] analysis of geographic variation in income inequality and intergenerational income mobility in the U.S. [26]. Color coded maps of geographic variation in prescription rates, overdose deaths, and economic factors suggested that regions of the country with declining upward income mobility tended to be regions that had higher opioid prescription and overdose rates [25, 26]. In support of the maps, our analyses [5] found that opioid prescriptions, social capital, and unemployment explained about 52% to 69% of the state-level variance in opioid overdose rates for the years 2001 to 2014. However, states are heterogeneous regarding the measures that we used, so that a more fine-grained analysis might reveal a more precise and nuanced account. In 2022, the CDC made county-level overdose data publicly available. This new data set included every U.S. county, including the many small ones whose overdose rates had heretofore been suppressed [27]. Thus, we were able to carry out the analyses presented in this report. The CDC also made county-level opioid prescription rate data available, starting in 2006 [28]. Hence, our analyses start with the year 2006.

## Materials and Methods

### The dependent variable

Our dependent variable was Midwest county drug overdose death rates from 2006 to 2020. However, in the interest of privacy, the CDC suppresses drug overdose data when the number of overdoses falls below 10. As most counties in the U.S. are sparsely populated, much of the county data are unavailable when the analyses are year by year. For example, for the year 2020, the CDC suppressed the overdose rates for 73% of Midwest counties. The CDC dealt with this issue by introducing an alternative, regression based, “model” overdose data set. According to the authors, this approach produced stable, yearly county overdose estimates that approximated the suppressed rates, although with a bias for underestimation [27].

Our tests supported their account. We aggregated the county results at the state level (which was not subject to suppression). The correlation between the regression-model, state overdose rates and the observed state overdose rates was *r*(178) = .97, and the median overdose rates for the two data sets—pooling across states and years—were 10.0 and 11.7 per 100000, respectively. Thus, the model overdose rates provide a valid account of the suppressed overdose rates. However, the model rates remained heavily right-skewed. To correct for this, we conducted our statistical analyses on the natural logs of the model rates. This reduced the skew indices to an acceptable range (from about a range of 1.55 to 2.30 to about a range of 0.18 to 0.36). Taking logs also simplified the interpretation of the results. Under this transformation, regression coefficients approximate the percentage change in the dependent variable associated with a one-unit change in the predictor [29].

### Predictors

The individual year regression models included seven predictors: intergenerational income mobility, opioid prescription rates, social capital, unemployment rate, student to teacher ratio, percentage of single parent families, and having attended college. We chose these predictors on the basis of our state-level analyses of opioid overdoses [5], which, in turn, were motivated by the overdose, prescription, and intergenerational income mobility maps, as described in the *Introduction*. The predictors were standardized, thereby facilitating comparisons across different predictors and analyses.

The *Opportunity Insights* research group, located at Harvard University, provided the county-level measures of intergenerational income mobility [30]. This measure is based on an adult child’s income rank relative to their parents’ income rank [31]. The parents’ income was set at its average percentile rank in 1994, 1995 and 1998-2000, where percentile rank is relative to the national income distribution of family incomes for those years. For example, a family in the 25^th^ percentile had an income that was higher than 24% of U.S. families. An adult child’s income percentile rank was measured relative to their birth cohort (1978 – 1983) in the years 2014 and 2015, when they were 31 to 37 years old. The data were obtained from national de-identified tax records. The *Opportunity Insights* analyses and publications focus on families whose 1990s’ income put them in the 25^th^ percentile rank—the midpoint of the lower half of the income distribution. We used this measure (*e_rank_b*), which is listed in Online Data Table 12 (“County-level Characteristics”) of the *Opportunity Insights* website. In words, it is the income percentile of individuals who were 31 to 37 years old in 2014 and 2015, and whose parents were in the 25^th^ income percentile in the 1990s.

The CDC provides county-level opioid prescription rates per 100 county residents [28]. They calculate the rates on the basis of pharmacy sales and Census Bureau population estimates. In the Midwest, approximately 90% of county pharmacies contributed data over the years 2006 to 2020, with a range of 85 to 98% depending on year. According to the CDC, failures to report sales were due to error or simply no sales. Because of this ambiguity, our analyses did not include counties that reported no sales. The opioid drugs were buprenorphine, codeine, fentanyl, hydrocodone, hydromorphone, methadone, morphine, oxycodone, oxymorphone, propoxyphene, tapentadol, and tramadol. The CDC’s prescription data date back to 2006, which is why our analyses start at 2006. The overdose rates, along with the social economic predictors, were downloaded in March of 2022 (see References).

In our earlier state-level analyses [5], social capital was the most robust socioeconomic predictor. Putnam [32] defined it as “features of social life—networks, norms, and trust—that allow participants to work together more effectively to pursue shared objectives.” As did Chetty and his colleagues (e.g., Data Table 12), we relied on Rupasingha et al.’s [24] operational index of Putnam’s definition, which has four components: associational densities (for example, membership rates in political organizations and professional organizations), the response rate for the Census Bureau’s population and housing survey, percentage of voters who voted in presidential elections, and per-capita non-profit organizations. As in our previous paper, we used social capital as measured in 1990. Unemployment rates were provided by the 2000 U.S. Census and were listed in *Opportunity Insights* Data Table 12. The Census Bureau defines unemployment as the percentage of individuals 16 years and older who were looking for work but not working during the week of the survey. County-level pupil/teacher ratio calculates the number of students per teacher in K through 12 public schools.

Chetty and colleagues used the data from the 1996-1997 Department of Education survey and listed the results in Data Table 12 of their website. The percentage of single parent families was defined as the number of households headed by a single mother that included one or more children under 18 years old, divided by the total number of households that included at least one child. The 2000 Census provided the data, which are listed in Data Table 10 of the *Opportunity Insights* website. “Some college” was defined as the percentage individuals in the 1978 to 1983 birth cohort who had at least some college experience by age 25 and was listed in Data Table 5. Thus, one education measure reflects elementary and high school experience, and the other reflects college experience and most predictors date from before the first year of our analyses (2006).

### Percent opioid overdose deaths

We calculated the percentage of overdose deaths that were associated with an opioid, using the CDC opioid categories T40.0 – T40.4 and T40.6 [1]. One calculation was based on just this list; the other was based on this list plus the category: “unspecified drugs.” Many coroners and medical examiners do not identify the drug associated with the death but instead simply note: “unspecified drug” [33, 34]. There is evidence that in many, if not most, cases, the “unspecified drug” was an opioid. For example, we found that opioid prescriptions were more strongly correlated with opioid plus “unspecified” overdose deaths than with opioid overdoses taken alone [5]. Consequently, we calculated the percentage of overdose deaths that were associated with opioids in two ways: with and without the “unspecified” instances. The denominator for these calculations was the CDC’s tally of all “drug poisonings.” All the data presented and discussed in this report are publicly available.

### Analytic approach

The yearly multilevel analyses proceeded in two-steps. First, we evaluated a random intercept model with no predictors (Model 1): *Y*_*ij*_ = *B*_00_ + *u*_0*j*_ + *e*_*ij*_, where *i* indicates county and *j* varies from 1 to 12 for state. *B*00 is the grand intercept, representing the overall overdose rate across all states, *u*_0*j*_ is the state-level residual, capturing the difference of each state’s intercept from the grand intercept, and *e*_*ij*_ is the county-level residual, capturing the between county, within state differences. On the basis of this model, we computed the proportion of variance in overdose deaths associated with between state differences, using the equation Var (*u*_0*j*_) /[Var(*u*_0*j*_) + *Var*(*e*_*ij*_)]. This quantity is referred to as the interclass correlation (ICC); the larger the ICC, the greater the influence of state-level differences on overdose deaths. For the years 2006 to 2020, the values ranged from 0.43 to 0.49, which confirmed the appropriateness of the multilevel approach. Next we included the 7 county-level predictors (Model 2—random intercept, fixed slope): *Y*_*ij*_ = *B*_00_ + *B*_10_ *x*1_*ij*_ + *B*_20_ *x*2_*ij*_ … + *B*_70_ *x*7_*ij*_ + *u*_0*j*_ + *e*_*ij*_, where x1 to x7 represent each of the seven predictors (e.g., prescription rates, social capital, etc.). In these models, a predictor’s slope was fixed, i.e., assumed to be the same in each state.

We also fit a model to the entire data set, pooling across years. This model included “year,” which was coded 1 to 15 (Model 3). For each model, we also computed the proportion of variance in overdose deaths accounted by the fixed effects of the predictors [35, 36].

## Results

### Prescription rates decrease, yet overdose rates continue to increase

Fig. 1 shows the average drug overdose rates per 100,000 individuals [27] and the average opioid prescription rates per 100 individuals [28] as a function of year. The overdose and prescription averages are based on county data, pooling across the 12 Midwest states. From 2006 to 2017, overdose rates steadily increased; in 2018 and 2019, they dipped slightly; then, in 2020, the increasing trend resumed. Overall, increases after 2013 were larger than increases prior to 2013. Although all states and most counties showed similar trends, there was much within and between state variability. The standard deviations, shown by the error bars, increased by about a factor of 2.7 between 2006 and 2020; the range (not shown), as measured by the 10^th^ and 90^th^ percentiles, increased by about a factor of 4.4 (7.89 and 34.77 deaths per 100,000). For the years 2006 to 2020, the highest average state overdose was 19.3/100000 (Ohio), and the lowest was 5.3/100000 (Nebraska).

**Fig 1:**
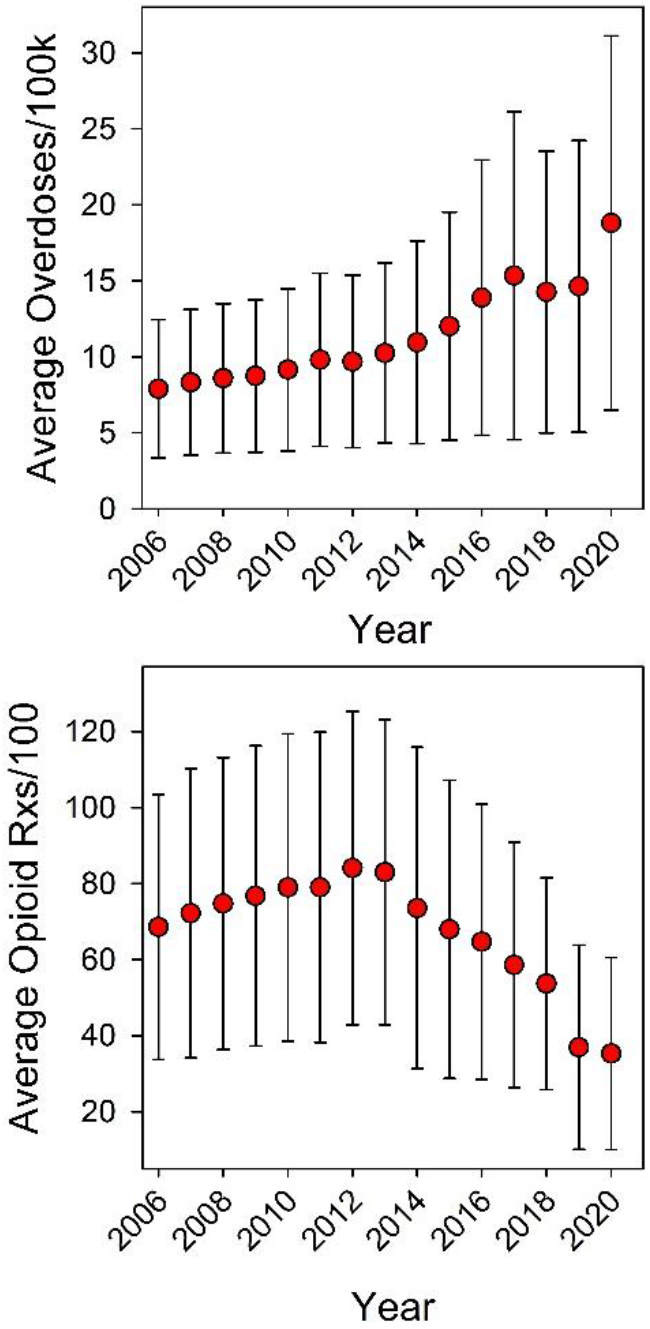
Midwest overdose and opioid prescription rates. Rossen et al. [27] of the CDC provided the overdose rates. The CDC Opioid Dispensing Rates Map site [28] provided the opioid prescription data. The error bars index the standard deviations for the 1055 Midwest counties.

Opioid prescription rates also increased from 2006 to 2012, but, then turned downward, decreasing each year. The decreases reversed previous increases, so that in 2019 and 2020, the prescription rates were lower than they were in 2006.

Fig. 2 shows the percentage of overdose deaths that were linked to an opioid as a function of year. The percentages were based on state-level data, pooling across states. As discussed in the Methods section, we calculated the percentage of opioid related overdoses in two ways: with and without “unspecified” overdose deaths. Either way, the percentage of opioid related deaths did not decrease even though legal opioid prescription sales decreased.

**Fig 2.**
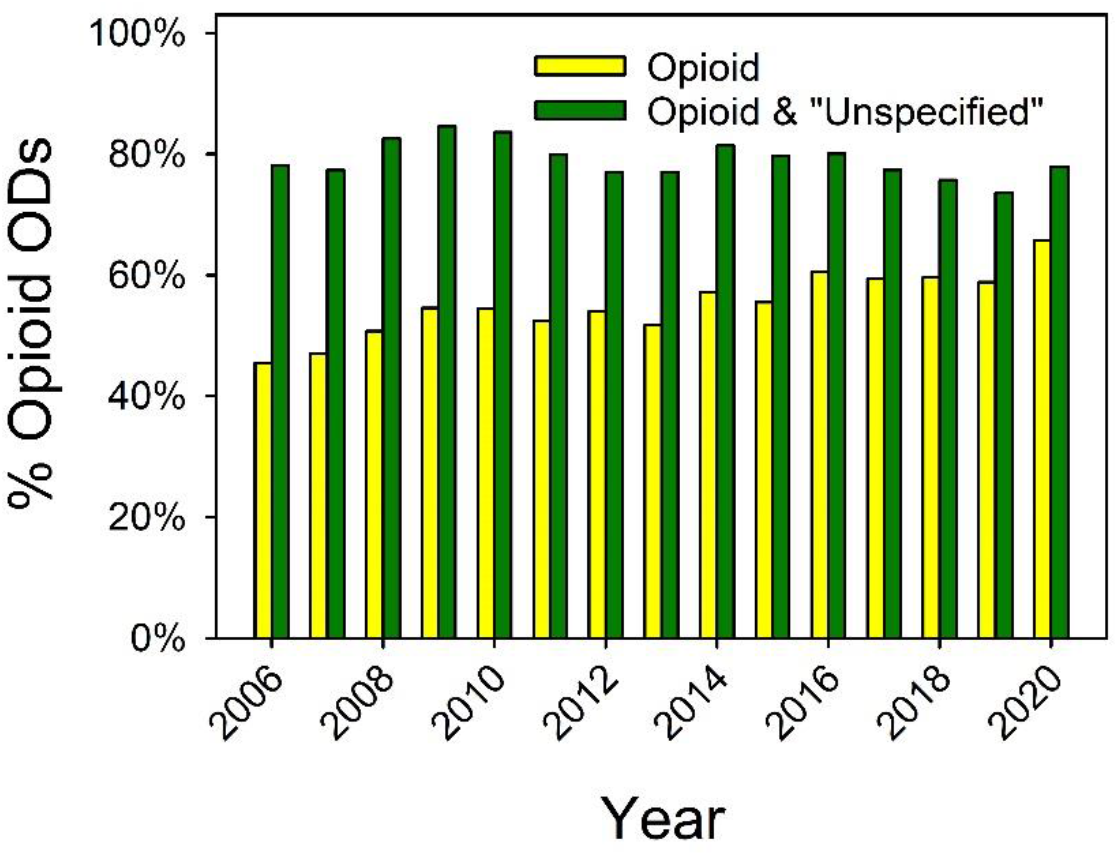
Percentage of opioid related overdose deaths. The CDC WONDER online data base [1] provided the data used to calculate opioid related overdoses. The *Methods* section provides the CDC codes for identifying opioids.

### Intergenerational income mobility was the strongest correlate and predictor of drug overdoses in Midwest counties

Table 1 lists the correlations between the natural logs of the overdose rates and the predictors for selected years. (See M*ethods* for a discussion of the advantages of running the statistical analyses on the logs of the overdose rates.) The correlations between the socioeconomic predictors and overdose rates varied from 0.46 to 0.68 (absolute values) and changed little as a function of year.

**Table 1.** Simple correlations for selected years among the (natural) log of the overdose rates and the predictors. *

|  | Log 2006<br>overdose<br>death | Log 2009<br>overdose<br>deaths | Log 2012<br>overdose<br>deaths | Log 2015<br>overdose<br>deaths | Log 2018<br>overdose<br>deaths | Log 2020<br>overdose<br>deaths |
| --- | --- | --- | --- | --- | --- | --- |
| Intergen. Income<br>Mobility | -0.68 | -0.67 | -0.68 | -0.68 | -0.68 | -0.68 |
| Social Capital | -0.54 | -0.55 | -0.54 | -0.52 | -0.53 | -0.51 |
| Unemployment<br>Rate | 0.52 | 0.53 | 0.51 | 0.51 | 0.51 | 0.50 |
| Pupil/Teacher<br>Ratio | 0.49 | 0.49 | 0.49 | 0.51 | 0.51 | 0.51 |
| % Single Parents | 0.47 | 0.47 | 0.46 | 0.47 | 0.46 | 0.47 |
| Attend College | -0.53 | -0.53 | -0.51 | -0.50 | -0.51 | -0.51 |
| Rx Rate 2006 | 0.45 | 0.45 | 0.44 | 0.43 | 0.43 | 0.43 |
| Rx Rate 2009 | 0.49 | 0.49 | 0.48 | 0.47 | 0.48 | 0.48 |
| Rx Rate 2012 | 0.45 | 0.46 | 0.44 | 0.43 | 0.43 | 0.43 |
| Rx Rate 2015 | 0.41 | 0.41 | 0.41 | 0.40 | 0.39 | 0.39 |
| Rx Rate 2018 | 0.32 | 0.32 | 0.31 | 0.29 | 0.29 | 0.29 |
| Rx Rate 2020 | 0.22 | 0.23 | 0.23 | 0.22 | 0.22 | 0.21 |

\* The CDC provided the overdose and prescription sales rates. The *Opportunities Insight* website provided the predictors. See *Methods* for details. All the correlations are statistically significant at the 0.05 level.

Intergenerational income mobility had the strongest correlation with overdoses, with a median value of – 0.68. In contrast, the year-to-year correlations between opioid prescriptions and overdoses had a more complex pattern. From 2006 to 2012, they ranged from about 0.44 to 0.49, but, then, steadily declined. For instance, by 2020, the correlation between opioid prescriptions and overdoses was about half as strong as it was in 2006. All the correlations were statistically significant at the <0.05 level. Table 2 provides a convenient summary of the year-to-year correlations. It is based on the pooled data.

**Table 2.**
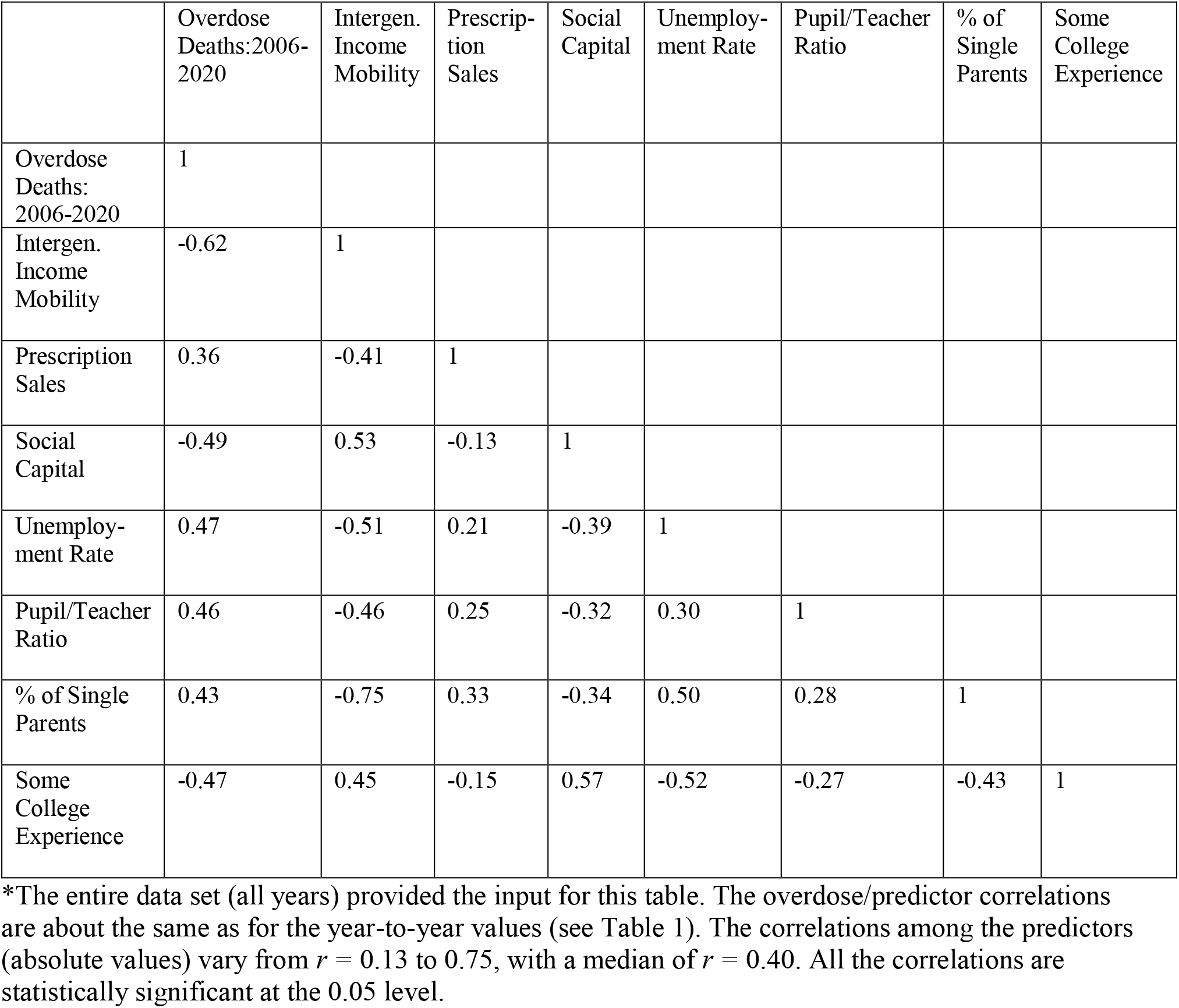
Pooled data correlations.*

Fig. 3 and Table 3 summarize the results for the multilevel regression analyses. The graph shows the regression coefficients for selected years (the same ones as in Table 1), and the table lists the coefficients for all years, along with the proportions of variance accounted for by the fixed effects. Since the units of the dependent variable are natural logs, and the units of the predictors are z-scores, the coefficients in Table 3 and Fig. 3 approximate the percentage difference in overdose rates for a one standard deviation difference in the predictor.

**Fig. 3:**
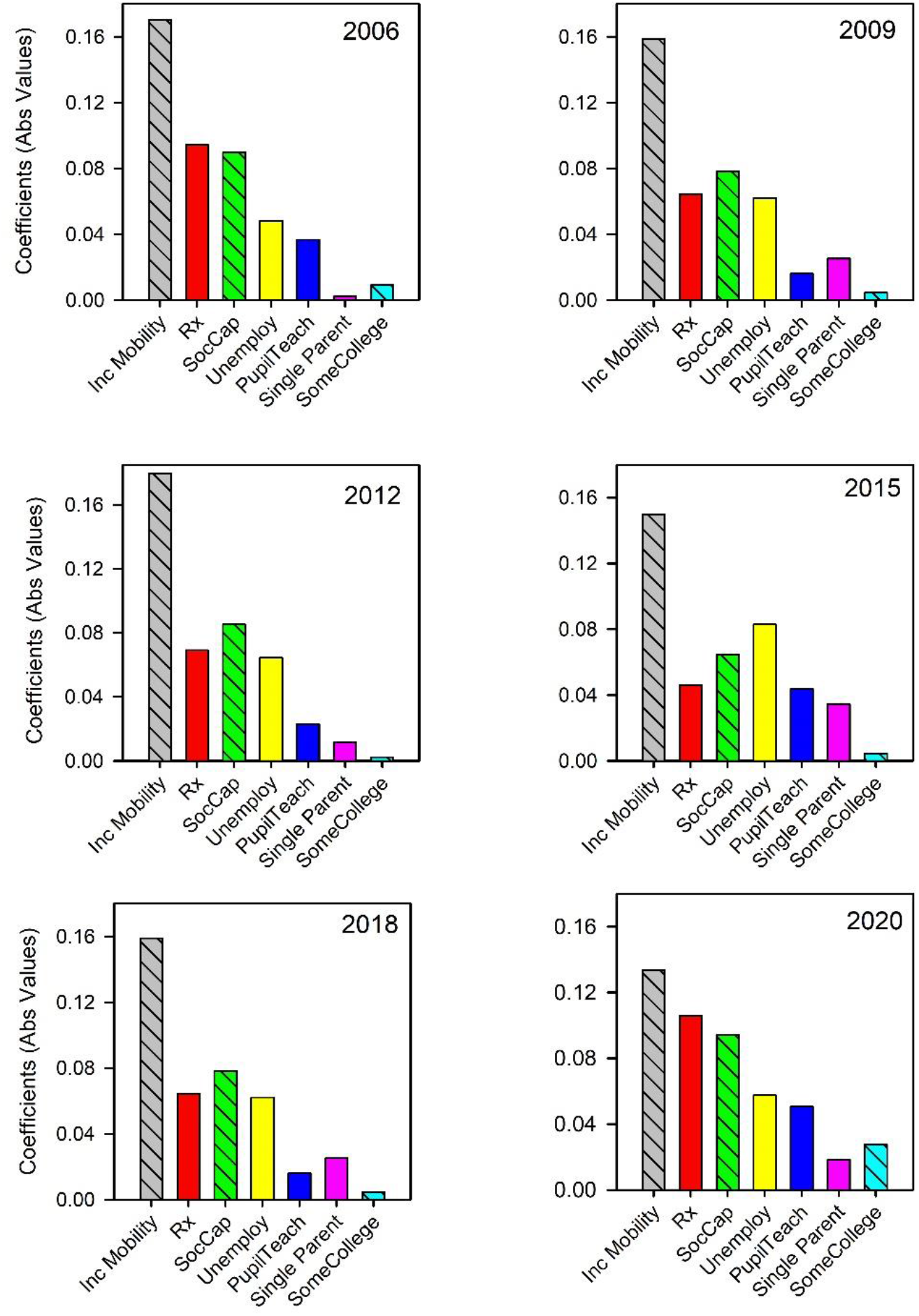
The fixed-effect regression coefficients for selected years. These are based on the yearly multilevel regressions. The negative diagonals indicate that the coefficient is negative, meaning the higher the value, the lower the overdose rate. Table 3 lists the numerical values and other statistical details for all years.

**Table 3.** Multilevel model results for each year of the study and the pooled data (Models 2 and 3).*

|  | Model 2* |  |  |  |  |  |  |  |  |  |  |  |  |  |  | Model 3 |
| --- | --- | --- | --- | --- | --- | --- | --- | --- | --- | --- | --- | --- | --- | --- | --- | --- |
| Year (n) | 2006 (870) | 2007 (865) | 2008 (866) | 2009 (865) | 2010 (863) | 2011 (864) | 2012 (861) | 2013 (862) | 2014 (929) | 2015 (930) | 2016 (931) | 2017 (926) | 2018 (903) | 2019 (945) | 2020 (944) | Pooled data (13,424) |
| *Fixed effects* |  |  |  |  |  |  |  |  |  |  |  |  |  |  |  |  |
| Intercept | 1.92 | 1.97 | 2.01 | 2.02 | 2.06 | 2.13 | 2.12 | 2.18 | 2.23 | 2.32 | 2.46 | 2.55 | 2.49 | 2.51 | 2.76 | 1.80 |
| Intergenerational Income Mobility | -0.143 | -0.142 | -0.156 | -0.134 | -0.143 | -0.144 | -0.170 | -0.167 | -0.159 | -0.159 | -0.139 | -0.167 | -0.180 | -0.159 | -0.149 | -0.144 |
| Opioid Rx Rate | 0.088 | 0.096 | 0.089 | 0.106 | 0.100 | 0.099 | 0.094 | 0.090 | 0.079 | 0.064 | 0.064 | 0.072 | 0.069 | 0.052 | 0.046 | 0.074 |
| Social Capital | -0.081 | -0.085 | -0.085 | -0.096 | -0.086 | -0.087 | -0.090 | -0.084 | -0.090 | -0.079 | -0.085 | -0.077 | -0.084 | -0.082 | -0.064 | -0.082 |
| Unemployment Rate | 0.064 | 0.060 | 0.060 | 0.057 | 0.055 | 0.051 | 0.049 | 0.047 | 0.060 | 0.061 | 0.050 | 0.060 | 0.065 | 0.067 | 0.083 | 0.058 |
| Pupil/Teacher Ratio | 0.043 | 0.045 | 0.048 | 0.050 | 0.054 | 0.049 | 0.036 | 0.044 | 0.022 | 0.016 | 0.018 | 0.008 | 0.023 | 0.037 | 0.043 | 0.031 |
| Single Parent | 0.016 | 0.013 | 0.007 | 0.019 | 0.012 | 0.007 | -0.003 | 0.000 | 0.009 | 0.025 | 0.033 | 0.024 | 0.011 | 0.028 | 0.034 | 0.025 |
| Some College | -0.018 | -0.023 | -0.024 | -0.027 | -0.015 | -0.019 | -0.009 | -0.004 | -0.002 | 0.005 | 0.010 | -0.012 | -0.003 | -0.003 | -0.004 | -0.005 |
| Year | --- | --- | --- | --- | --- | --- | ---- | -- | --- | --- | --- | --- | --- | --- | --- | 0.055 |
| *Random effects* |  |  |  |  |  |  |  |  |  |  |  |  |  |  |  |  |
| Var(u_0j) | .036 | .035 | .032 | .029 | .038 | .037 | .039 | .044 | .049 | .056 | .072 | .069 | .057 | .057 | .062 | .053 |
| Var(e_ij) | .091 | .091 | .092 | .092 | .095 | .093 | .097 | .094 | .100 | .103 | .106 | .112 | .103 | .106 | .105 | .103 |
| Explained variance | 0.43 | 0.44 | 0.46 | 0.47 | 0.43 | 0.43 | 0.42 | 0.40 | 0.38 | 0.34 | 0.29 | 0.33 | 0.37 | 0.35 | 0.34 | 0.47 |

Each year, intergenerational income mobility was the strongest predictor of drug overdose deaths. From 2006 to 2009, prescription rates were the second strongest predictor, but then, as suggested by Figure 1 and Table 1, the association with prescriptions steadily declined. For instance, in 2018 opioid prescriptions were the fourth strongest predictor, and in 2020, they were the fifth strongest. On average, the regression coefficients for income mobility were 1.91 times larger than the regression coefficients for prescription rates.

Social capital was the second strongest predictor and, slightly behind it, prescription rates were the third strongest predictor. Having gone to college for a year or more was the weakest predictor. Put in terms of average percentage change, a one standard deviation increase in intergenerational income mobility and social capital was associated with a 14.5% and 8.3% decrease in overdose deaths. In contrast, a one standard deviation increase in opioid prescription sales and unemployment rates was associated with a 7.4% and 5.8% increase in overdose deaths. As measured by the variance accounted for proportion, the predictive strength of the regression models was usually above 40%.

Fig. 4 provides a representative summary of the regression results, collapsing across all years. On the y-axis are the regression coefficients for the pooled data. As noted in the *Materials and Methods* section, this model included year as a predictor, which was coded 1 to 15. The coefficients for the pooled data are will within the range of those obtained in the year-by-year analyses. Again, intergenerational income mobility was the strongest predictor of overdose deaths, and again the coefficient for income mobility was about twice as large as the coefficient for opioid prescriptions. This model accounted for 47% of the variance in overdose deaths, pooling across all years and counties.

**Fig. 4.**
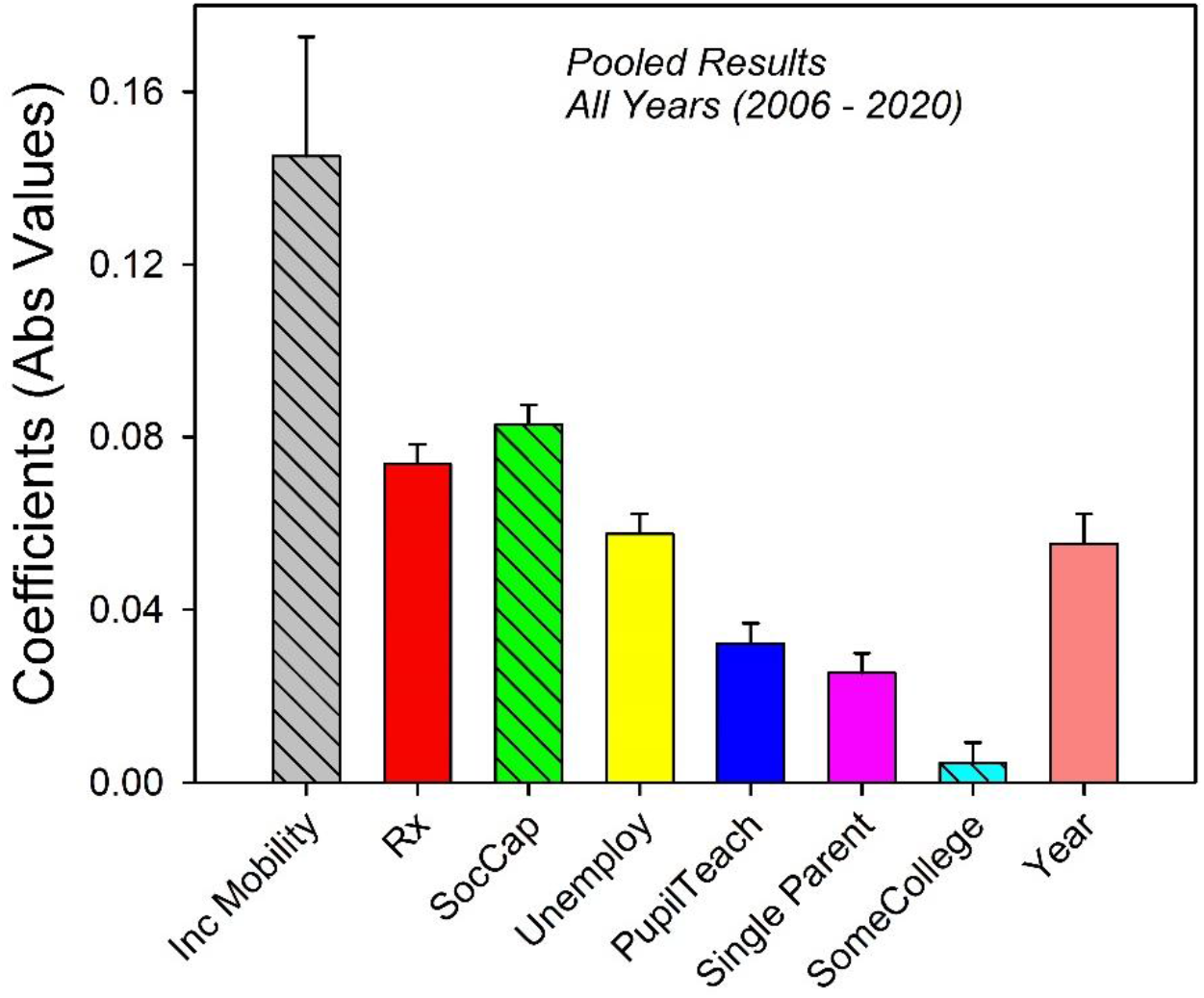
The fixed effects regression coefficients for the pooled data for all years from 2006 to 2020. This analysis included year as an 8^th^ predictor. It was coded 1 to 15. Otherwise the format is the same as that for Fig. 3.

## Discussion

### Summary

In 2020 approximately 21% of the population of the U.S. lived in the 1055 counties of the 12 Midwest states. In these counties, the strongest predictor of overdose deaths was the income rank of adult children relative to their parent’s income rank: the lower the rank, the higher the overdose rate. The regression coefficients linking intergenerational mobility (or lack thereof) and overdose deaths were about twice as large as those linking opioid prescription rates to overdose deaths. The second largest, albeit by a slim margin, were those for social capital. The third largest were those for opioid prescriptions. As described next, these results are consistent with the framework outlined in the beginning of this report: the large influx of prescription opioids in the years 1995 to 2012 and the concomitant increase in overdose deaths are episodes in a larger story.

### Intergenerational income mobility provides a socioeconomic context for four decades of rising drug overdose rates

First, Figure 1 implies that illegal opioid and non-opioid drugs replaced prescription opioids as the sources for most drug overdose deaths. In support of this point, Ruhm [37] found that opioid and non-opioid overdose deaths occurred at about the same rate for the years that relevant data were available (1999 – 2016). More tellingly, Jalal and colleagues (2018) reported that from 1979 to 2016 (the years over which data were collected), U.S. drug overdose deaths increased at a steady, slightly increasing, exponential rate—apart from a bump in the rate of increase from about 2004 to 2010. The particular drugs that were most toxic changed, but the overall trend, counting all drugs, did not.

Importantly, this trend started well before OxyContin and other prescription opioids became widely available, and, likewise, it has continued at about the same rate after prescriptions were less available. Thus, as measured by rate, opioid prescription overdose deaths are continuous with earlier and later periods of drug overdose deaths. (The former involved stimulants, the latter, including up to the present, involve fentanyl and heroin.)

Second, if drug availability and social-conditions jointly determine large, societal-level changes in drug use (as noted in the *Introduction*), there should exist social factors that parallel the steady increase in drug overdose deaths that date to 1978 and possibly earlier. The regression analyses say that one such factor, and possibly the most important one, is the decrease in upward intergenerational income mobility. Historical trends in income mobility support this hypothesis.

Chetty et al. [38] measured changes in intergenerational income mobility for cohorts born between 1940 and 1984 for the entire U.S. These cohorts entered their 30s and 40s in just the years that drug overdose rates embarked on their current increasing trend (1978 to the present). For the years in question, intergenerational income mobility steadily decreased. The 1940 birth cohort had more than a 90% likelihood of reaching a higher income rank than their parents; whereas, individuals born in 1984 had a 50% likelihood of reaching a higher income rank than their parents. Thus, overdoses deaths have been steadily increasing just as relative economic prospects have been steadily declining. Chetty et al.’s analysis is based on tens of millions of individual tax records, drawn from every state and county in the country; our data are Midwest county records; both support the same conclusion: intergenerational income mobility is a strong, if not the strongest, socioeconomic predictor of drug overdose trends in the United States.

Jalal and colleagues [19, p. 5] point out that “understanding the forces” associated with the “multiple subepidemics” that make up the now nearly half-century long rise in drug overdose deaths may help reveal the “root causes of the epidemic” and this understanding “may be crucial to … prevention and intervention strategies.” Our data suggest that intergenerational income mobility is a strong “root cause” candidate.

### Advantages and limitations

We selected the Midwest states on the basis of data suggesting that trends in the overdose data would be simpler to interpret for this region of the country. Previous state and county-level analyses show that overdose rates as well as their predictors differ according to race and ethnicity [5, 39]. The model overdose data used in this report are based on the entire population. Thus, it is possible that the advantages that this data set provided (e.g., stable year-to year overdose rates in sparsely populated counties) would be offset by group differences. However, in the 12 Midwest states, overdose rates are similar for Whites and Non-Whites (17.3 and 17.0/100k). This does not remove the potential problem posed by samples that include heterogeneous populations, but it is consistent with the assumption that the predictors of overdose deaths for different racial and ethnic groups are more similar in the Midwest than in other regions of the country. (Group differences in the predictors of overdose deaths are not well understood. For comments on this topic at the county and state levels, see Cano et al. [39] and Heyman et al. [5]).

Of course, this advantage comes with a potential disadvantage. Similar analyses for other regions of the country may yield more complex accounts of the predictors of overdose rates. However, the results summarized in this report proved consistent with national analyses of overdose rates that included social capital and family structure as predictors [5, 13, 23], and, as emphasized, the Midwest results are consistent with the nation-wide correlation between decreasing upward economic mobility and increasing overdose deaths.

A second limitation is in regards to the factors that mediate the association between socioeconomic predictors and overdose rates. Individuals—not counties—purchase and consume drugs. Case and Deaton’s [40] often referenced claim that Americans have entered an era of “despair” suggests hypotheses as to the mediating psychological factors. Taking up this challenge, Dasgupta and colleagues [12] proposed that the tendency to somaticize socioeconomic frustrations connects despair to opioids. However, to our knowledge, research has yet to flesh out this possible link.

## Conclusion

Drug availability has hitherto been a powerful correlate of drug use and addiction. For soldiers stationed in Vietnam in the 1960s and 1970s, the wide availability of opiates was accompanied by marked increases in their use [41]. However, when these men returned to the U.S., metabolic tests revealed that regular use, along with signs of dependence, declined by almost 90%. In contrast, the decrease in the availability of legal opioid pain-killers, which started in 2013, has not been followed by a decrease in drug overdose deaths, even though the bulk of such deaths were associated with opioid use. The simplest explanation is that (1) we are in an era in which illegal drugs are relatively easy to come by, and (2) the steady decrease in upwards intergenerational income mobility has fueled a steady increase in the pool of individuals attracted to the intoxicating effects of toxic drugs.

## Data Availability

1. cdc.gov [Internet]. Atlanta (GA): CDC Wonder: Multiple Cause of Death, 19992020 Request; [cited 2023 June 27]. Available from: https://wonder.cdc.gov/mcdicd10.html

2. usdoj.gov [Internet]. Arlington (VA): United States Department of Justice Drug Enforcement Administration; [cited 2023 June 27]. Available from: https://www.deadiversion.usdoj.gov/arcos/retaildrugsummary

18. stacks.cdc.gov [Internet]. Atlanta (GA): National Center for Health Statistics; [cited 2023 Jun 28]. Available from: http://doi.org/10.15620/cdc:112340

27. Rossen LM, Bastian B, Warner M, Khan D, Chong Y. County-level drug overdose mortality: United States, 20032020 [Data file and Map]. National Center for Health Statistics: Hyattsville(MD); 2022. [cited 2023 Jun 28]. Available from: https://www.cdc.gov/nchs/datavisualization/drugpoisoningmortality/#datatables

28. CDC Injury Center [Internet]. US: The Center; 2006-2020 [cited 2023 Jun 28]. Opioid Dispensing Rate Maps; [about 5 screens]. Available from: https://www.cdc.gov/drugoverdose/rxratemaps/index.html

30. opportunityinsights.org [Internet]. Cambridge: Harvard University; c2023 [cited 2023 Jun 28]. Available from: https://opportunityinsights.org

https://wonder.cdc.gov/mcd-icd10.html

https://www.deadiversion.usdoj.gov/arcos/retail_drug_summary

http://doi.org/10.15620/cdc:112340

https://www.cdc.gov/nchs/data-visualization/drug-poisoning-mortality/#data-tables

https://www.cdc.gov/drugoverdose/rxrate-maps/index.html

https://opportunityinsights.org

## Acknowledgments

We appreciate the help of Zane Madi, Nikul Patel, and Liuying Huang in preparing this manuscript. Requests for materials should be sent to Gene Heyman. Funding: The research was supported by funds from Boston College.

## Data and materials availability

All data are publicly available. The websites that provided the data are listed in text and *References*.

## Author Contributions

Conceptualization: GMH, HB, ER Data Curation: GMH

Formal Analysis: ER, GMH, HB Investigation: GMH Methodology: ER, GMH, HB Validation: GMH

Writing: GMH

Writing, Review, Editing: HB, ER, GMH

## References

1. cdc.gov [Internet]. Atlanta (GA): CDC Wonder: Multiple Cause of Death, 1999-2020 Request; [cited 2023 June 27]. Available from: https://wonder.cdc.gov/mcd-icd10.html

2. usdoj.gov [Internet]. Arlington (VA): United States Department of Justice Drug Enforcement Administration; [cited 2023 June 27]. Available from: https://www.deadiversion.usdoj.gov/arcos/retail_drug_summary.

3. Bohnert ASB, Ilgen MA, Trafton JA, Kerns RD, Eisenberg A, Ganoczy D, et al. Trends and regional variation in opioid overdose mortality among Veterans Health Administration patients, fiscal year 2001 to 2009. Clin J Pain. 2014 Jul;30(7):605–12. doi: 10.1097/AJP.0000000000000011. PMID: 24281278.

4. Griffith KN, Feyman Y, Auty SG, Crable EL, Levengood TW. Implications of county-level variation in U.S. opioid distribution. Drug Alcohol Depend. 2021 Feb;219:108501. doi: 10.1016/j.drugalcdep.2020.108501. PMID: 33421805.

5. Heyman GM, McVicar N, Brownell H. Evidence that socioeconomic factors play an important role in drug overdose deaths. Int J Drug Policy. 2019 Dec;74:274–84. doi: 10.1016/j.drugpo.2019.07.026. Epub 2019/8/28. PMID: 31471008.

6. Ghertner R. U.S. county prevalence of retail prescription opioid sales and opioid-related hospitalizations from 2011 to 2014. Drug Alcohol Depend. 2019 Jan;194:330–35. doi: 10.1016/j.drugalcdep.2018.10.031

7. Han B, Compton WM, Blanco C, Crane E, Lee J, Jones CM. Prescription opioid use, misuse, and use disorders in US adults: 2015 National Survey on Drug Use and Health. Ann Intern Med. 2017 Sep;167(5):293–301. doi: 10.7326/M17-0865. Epub 2017/8/1. PMID: 28761945.

8. Madras BK. The President’s Commission on combating drug addiction and the opioid crisis: origins and recommendations. Clin Pharmacol Ther. 2018 Jun;103(6): 943–5. doi: 10.1002/cpt.1050. Epub 2018/3/23. PMID: 29570781.

9. Meier B. Origins of an epidemic: Purdue Pharma knew its opioids were widely abused. The New York Times. 2018 May 29 [cited 2023 June 28]. Available from: https://www.nytimes.com/2018/05/29/health/purdue-opioids-oxycontin.html

10. Zhang S. The one-paragraph letter from 1980 that fueled the opioid crisis. The Atlantic. 2017 June 2 [cited 2023 June 28]. Available from: https://www.theatlantic.com/health/archive/2017/06/nejm-letter-opioids/528840/

11. Dave D, Deza M, Horn B. Prescription drug monitoring programs, opioid abuse, and crime. Southern Economic Journal. 2021 Jan;87(3):808–48. doi: 10.1002/soej.12481

12. Dasgupta N, Beletsky L, Ciccarone D. Opioid crisis: No easy fix to its social and economic determinants. Am J Public Health. 2018 Feb;108(2):182–6. doi: 10.2105/AJPH.2017.304187. PMID: 29267060.

13. Monnat SM. Factors associated with county-level differences in U.S. drug-related mortality rates. Am J Prev Med. 2018 May;54(5): 611–9. doi: 10.1016/j.amepre.2018.01.040. PMID: 29598858.

14. Szalavitz M. The social life of opioids. Scientific American. 2017 September 18 [cited 2023 June 27]. Available from: https://www.scientificamerican.com/article/the-social-life-of-opioids/

15. Quinones S. Dreamland: The true tale of America’s opiate epidemic. 1st ed. New York: Bloomsbury Press; 2015.

16. Quinones S. Raymond Sackler: the philanthropist who helped spawn the Opioid Crisis. POLITICO Magazine. 2017 December 28 [cited 2023 June 27]. Available from: https://www.politico.com/magazine/story/2017/12/28/raymond-sackler-obituary-216185/

17. Ciccarone D. The triple wave epidemic: supply and demand drivers of the US opioid overdose crisis. Int J Drug Policy. 2019 Sep;71:183. doi: 10.1016/j.drugpo.2019.01.010. Epub 2019/2/2. PMID: 30718120.

19. Jalal H, Buchanich JM, Roberts MS, Balmert LC, Zhang K, Burke DS. Changing dynamics of the drug overdose epidemic in the United States from 1979 through 2016. Science. 2018 Sep;361(6408): eaau1184. doi: 10.1126/science.aau1184. PMID: 30237320

20. Bailey P. The heroin habit. New Republic. 1916;6:314–6.

21. Courtwright DT. Dark paradise: A history of opiate addiction in America. Cambridge, MA: Harvard University Press; 2001.

22. Ruhm CJ. Drivers of the fatal drug epidemic. J Health Econ. 2019 Mar;64:25–42. doi: 10.1016/j.jhealeco.2019.01.001. PMID: 30784811.

23. Zoorob MJ, Salemi JL. Bowling alone, dying together: The role of social capital in mitigating the drug overdose epidemic in the United States. Drug Alcohol Depend. 2017 Apr;173:1–9. doi: 10.1016/j.drugalcdep.2016.12.011. PMID: 28182980.

24. Rupasingha A, Goetz SJ, Freshwater D. The production of social capital in US counties. Journal of Socio-Economics. 2006 Feb;35(1):83–101. doi: 10.1016/j.socec.2005.11.001

25. Chetty R, Hendren N, Kline P, Saez E. Where is the land of opportunity? The geography of intergenerational mobility in the United States. The Quarterly Journal of Economics. 2014 Jun;129(4): 1553–623. doi: 10.3386/w19843

26. McDonald DC, Carlson K, Izrael D. Geographic variation in opioid prescribing in the U.S. J Pain. 2012 Oct;13(10): 988–96. doi: 10.1016/j.jpain.2012.07.007. PMID: 23031398.

27. Rossen LM, Bastian B, Warner M, Khan D, Chong Y. County-level drug overdose mortality: United States, 2003–2020 [Data file and Map]. National Center for Health Statistics: Hyattsville(MD); 2022. [cited 2023 Jun 28]. Available from: https://www.cdc.gov/nchs/data-visualization/drug-poisoning-mortality/#data-tables

28. CDC Injury Center [Internet]. US: The Center; 2006-2020 [cited 2023 Jun 28]. Opioid Dispensing Rate Maps; [about 5 screens]. Available from: https://www.cdc.gov/drugoverdose/rxrate-maps/index.html

29. Wooldridge JM. Introductory Econometrics: A Modern Approach. 6th ed. Mason: Cengage Learning; 2015.

30. opportunityinsights.org [Internet]. Cambridge: Harvard University; 2023 [cited 2023 Jun 28]. Available from: https://opportunityinsights.org.

31. Chetty R, Hendren N, Jones MR, Porter SR. Race and economic opportunity in the United States: An intergenerational perspective. The Quarterly Journal of Economics. 2020 May;135(2):711–83. doi: 10.1093/qje/qjz042.

32. Putnam RD. Tuning in, tuning out: the strange disappearance of social capital in America. PS: Political Science and Politics. 1995;28(4):664–83. doi: 10.2307/420517.

33. Warner M, Paulozzi LJ, Nolte KB, Davis GG, Nelson LS. State variation in certifying manner of death and drugs involved in drug intoxication deaths. Acad Forensic Pathol. 2013 Jun;3(2):231–37. doi: 10.23907/2013.029.

34. Webster LR, Dasgupta N. Obtaining adequate data to determine causes of opioid-related overdose deaths. Pain Med. 2011 Jun;12(Supplement 2):S86–92. doi: 10.1111/j.1526-4637.2011.01132.x. PMID: 21668762

35. Rights JD, Sterba SK. Quantifying explained variance in multilevel models: An integrative framework for defining r-squared measures. Psychol Methods. 2019 Jun;24(3):309–38. doi: 10.1037/met0000184. PMID: 29999378.

36. Rights JD, Sterba SK. Effect size measures for longitudinal growth analyses: Extending a framework of multilevel model r-squared to accommodate heteroscedasticity, autocorrelation, nonlinearity, and alternative centering strategies. New Dir Child Adolesc Dev. 2021 Jan;2021(175):65–110. doi: 10.1002/cad.20387. PMID: 33512773

37. Ruhm CJ. Nonopioid overdose death rates rose almost as fast as those involving opioids, 1999–2016. Health Aff (Millwood). 2019 Jul;38(7):1216–24. doi: 10.1377/hlthaff.2018.05522. PMID: 31260365.

38. Chetty R, Grusky D, Hell M, Hendren N, Manduca R, Narang J. The fading American dream: Trends in absolute income mobility since 1940. Science. 2017 Apr;356(6336):398–406. doi: 10.1126/science.aal4617.

39. Cano M, Oh S, Osborn P, Olowolaju SA, Sanchez A, Kim Y, Moreno AC. County-level predictors of US drug overdose mortality: a systematic review. Drug Alcohol Depend. 2023 Jan 1;242:109714. doi: 10.1016/j.drugalcdep.2022.109714. Epub 2022 Nov 24. PMID: 36463764.

40. Case A, Deaton A. Rising morbidity and mortality in midlife among white non-Hispanic Americans in the 21st century. Proc Natl Acad Sci U S A. 2015 Dec;112(49):15078–83. doi: 10.1073/pnas.1518393112. Epub 2015 Nov 2. PMID: 26575631.

41. Robins LN. The sixth Thomas James Okey Memorial Lecture. Vietnam veterans’ rapid recovery from heroin addiction: a fluke or normal expectation? Addiction. 1993 Aug;88(8):1041–54. doi: 10.1111/j.1360-0443.1993.tb02123.x. PMID: 8401158.

